# Effect of Seating Arrangement on Class Engagement in Team-based Learning: A Quasi-Experimental Study

**DOI:** 10.1101/2020.10.25.20218313

**Authors:** Seet Hong An Andrew, Emmanuel Tan, Preman Rajalingam

## Abstract

**Introduction:** This study investigated the effects of seating distance and orientation on engagement in novice and experienced learners in a large classroom explicitly designed for Team-based Learning (TBL). Learning what affects TBL engagement may improve its implementation.

**Methods:** Participants were novice first-year and experienced second-year undergraduate medical students in Singapore (male=103, female=57). Their age ranged from 18 to 23 (M=19.5, SD=1.06). This quasi-experimental study considered two factors. Firstly, the distance from the teams’ table to the tutor’s table. Secondly, students’ orientation at each table, with either their front or back facing the tutor. Engagement was measured using two instruments, Situational Cognitive Engagement Measure and Classroom Engagement Survey at two TBL sessions – before and after swapping seating arrangements.

**Results:** For experienced students, seating distance did not significantly affect engagement (p=0.08–0.89). Novice student’s engagement levels decreased significantly for those who moved further; M=3.30 to 2.98 (p=0.009–0.023). However, overall engagement also decreased post-swap regardless of direction moved; M=3.26 to 3.00 (p=0.004). For both cohorts, seating orientation did not significantly affect engagement (p=0.07–0.62). Those unaffected by seating arrangement commended the classroom’s design, such as screens all around and quality audio-visual system. Novice students exhibited a stronger preference to sit nearer to the tutor than experienced students. Both groups preferred sitting with their front-facing the tutor.

**Discussion:** Within specially designed TBL classrooms, seating distance and orientation did not significantly affect engagement. Technologically enhanced team-centric spaces provide a favorable environment for TBL, though students’ preferences for seats may change with more TBL experience.

## Introduction

### Seating Arrangement and Engagement

There is a growing literature on the importance of learning spaces and how it affects teaching and learning[1]. Especially, learning spaces affecting the delivery of higher education[2] and the medical curriculum[3]. The effects of physical space on human activity has been studied where the area of environmental psychology examines subjects such as psychological comfort with space and its incentivising effects[4]. Classroom designs can vary in many forms to strengthen classroom discussions[5]. In traditional lecture hall settings, engagement is worst towards the back of the class[6], grades decreased as distance from the instructor increased, and a smaller proportion of students participated in larger class sizes[7]. On the other hand, semicircular arrangements allow for more comfortable discussion between tutor and students and within students themselves[8]. The students also asked more questions and learnt more compared to traditional row and column organization[9]. Such decentralised layouts encourage participation and show that the classroom’s physical attributes clearly affect the engagement of students.

Cognitive engagement is the psychological construct characterized by active involvement in learning, where students have autonomy[10] and put effort to invest in studying[11-13]. It varies depending on the activity[14], increasing as knowledge is gained, and with more independence in the task involved[15]. It is important in the learning process as cognitive engagement is related to positive outcomes in students, such as improved grades[16], motivation[17] and favorable attitudes towards learning[18].

### Team-Based Learning

Team-based learning (TBL) is a structured, learner-centered and instructor-led method of active learning relying on small group interaction and class-wide discussion[19, 20]. TBL use is increasingly popular in medical education[21] with positive outcomes in many areas from attitudes and graded assignments[22], to critical thinking, teamwork and communication[23, 24].

A TBL session comprises of three phases: preparation, readiness assurance and application exercise (AE) phase[25] (Figure 1). The preparation phase involves individual learning outside the classroom using resources provided by the school. The subsequent phases occur in the classroom. Firstly, the students start with an Individual Readiness Assurance Test, a closed book multiple-choice question test, which is then repeated in teams, the Team Readiness Assurance Test, and a “burning questions” (BQ) discussion. Lastly, the AE phase challenges students with practical scenarios, which provides them with an opportunity to apply their knowledge gained. This study was conducted at the Lee Kong Chian School of Medicine, Nanyang Technological University, Singapore. TBL is our main pedagogical method for the first two years of medical undergraduate learning. (Refer to Rajalingam et al[26], large-scale TBL paper for more information. TBL procedures described here may vary between institutions.)

**Figure 1:**
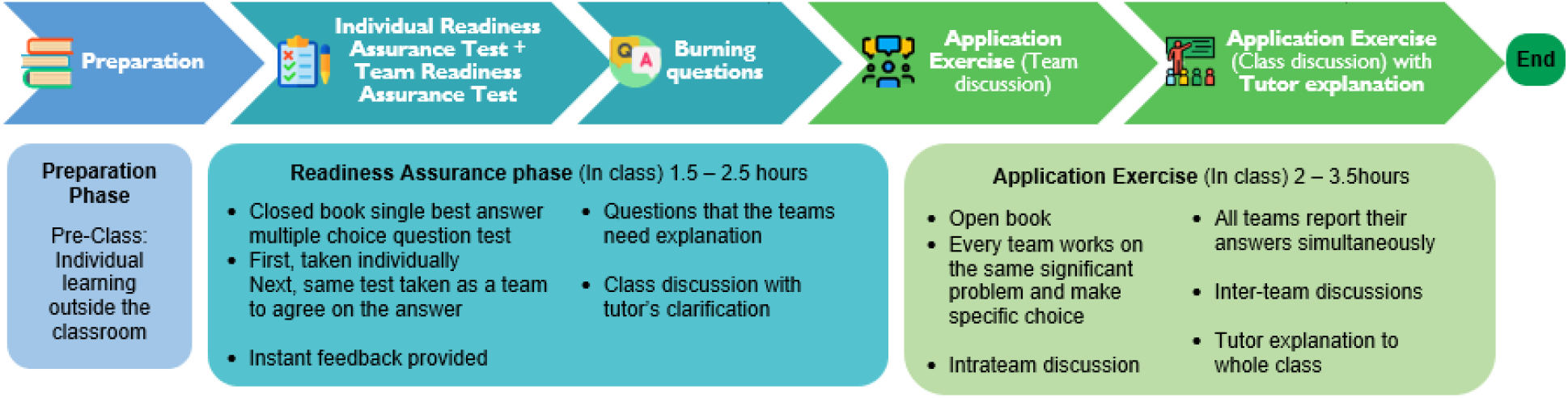
Schematic of a typical Team-Based Learning (TBL) process from the Preparation to Readiness Assurance, Burning questions and Application Exercise phase at the Lee Kong Chian School of Medicine, Nanyang Technological University, Singapore. Descriptions about what each phase entails and their average duration are included. TBL procedures described here may vary between institutions.

A theory about the underlying psychological process[27, 28] through which TBL works is that it increases students’ cognitive engagement and subsequently affects achievement[14].

Previous TBL research found a significant increase in students’ engagement[29-32], where the nature of the tasks promotes participation and discussion while making students process the information[25]. Thus, learning what affects engagement may improve the effective implementation of TBL.

Many medical schools that have implemented TBL have also made physical changes to the classroom to accommodate this instructional approach. In this study, our main interest is the effect of seating arrangement on TBL engagement. However, limited research exists in this field. One study by Espey M[33] examined classroom layouts and their impact on TBL. Although grades were not significantly affected, students felt they worked more effectively in classrooms that easily accommodate group work and attitudes improved. With varying cohort sizes among institutions[34], there may be concerns of large-scale TBL implementation as class size increases, students sitting further away maybe disadvantaged. Furthermore, in a previous local study, cognitive engagement fell significantly during BQ and AE tutor explanation phases compared to the previous phases[14]. Exploring the effects of seating arrangement may promote engagement during these tutor-centred phases. While other factors like gender have been examined in the context of seating position[35], the effect of exposure to TBL has not been explored, such as the differences between novice and experienced students.

In this study, we aimed to investigate two factors associated with seating arrangement. Firstly, the effects of seating distance from the tutor on class engagement in TBL. Secondly, if seating orientation within the team, which is the direction students are facing, affects class engagement in TBL. The hypotheses are: (1) sitting nearer to the tutor results in higher engagement and (2) sitting with their front facing the tutor class results in higher engagement.

## Methods

### Study Design

In this quasi-experimental study, students were assigned to teams of 5-7 based on TBL principles[36]. Our classroom design includes a dual-tiered seating circular layout, six big screens all around the periphery of the room, chairs with wheels around fixed tables and microphones for every team. Usually, the teams’ seating positions are permanent for the entire academic year. Tutors were situated in front and remained static for the purposes of this study, reflecting the default situation during TBL sessions. As teams were seated in a circular arrangement, seating orientation was reported as either sitting with their front or back to the tutor. Seating distance from the teams’ table to the tutor’s table ranged from 2.5m to 11m.

Engagement was assessed at two points: BQ and AE, both after tutor explanation. These were chosen as engagement fell significantly compared to the previous phases[14]. Also, seating distance likely affects engagement when the tutor speaks in front of the class, rather than during intra-team discussion at other phases where students discuss within their teams.

The intervention involved rearranging the students’ seating layout and collecting data before and after this swap (see Supplementary Digital Appendix 1 for lesson topics). After obtaining baseline response, teams were swapped front-to-back, with an average change of 4.5m. Students could choose their own seats at the new table. 12-13 teams moved nearer to the tutor, labelled as Group A while 11-12 teams moved further, labelled as Group B (Figure 2). One TBL session was given for students to adjust to their new positions before collecting responses at the subsequent lesson a week later. Researchers were present to conduct the surveys at appropriate intervals. We used SurveyMonkey platform to collect responses electronically.

**Figure 2:**
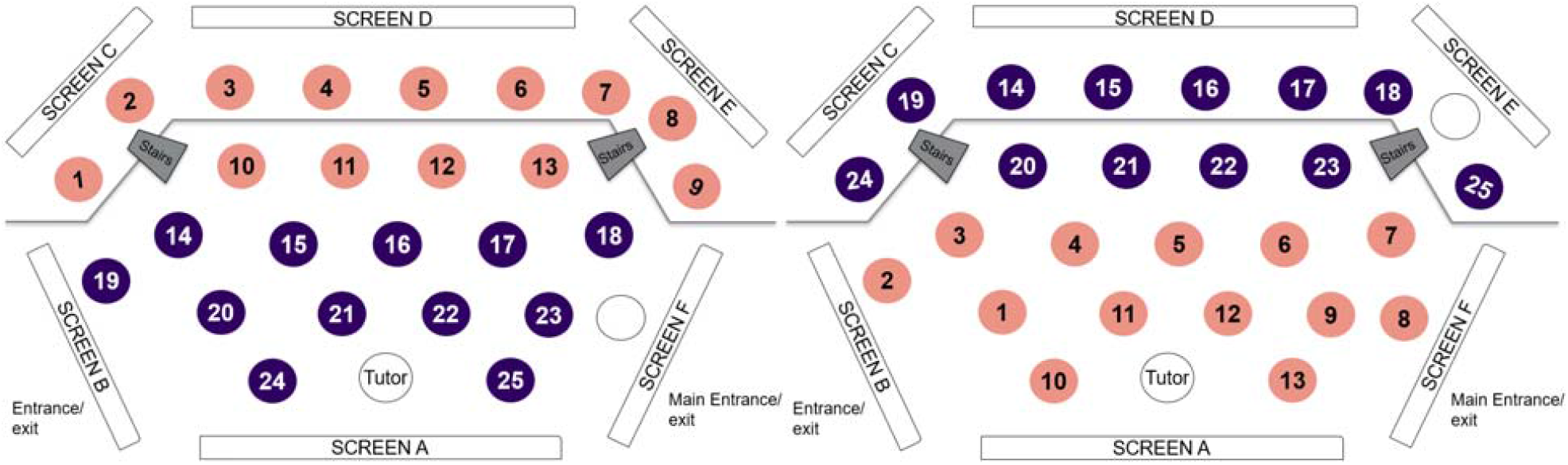
Classroom seating arrangement at the Lee Kong Chian School of Medicine, Nanyang Technological University, Singapore. The team numbers are represented by the circles. **Left**: Original seating positions before the swap. **Right**: New seating positions after swap. **Group A**: 13 teams (in orange) moved nearer. **Group B**: 12 teams (in blue) moved further.

There is a dual-tiered circular seating arrangement with 6 large screens all around the periphery, chairs with wheels around fixed tables and microphones for every team. Tutors are in front of the class and remained static.

### Participants

150 first-year and 138 second-year undergraduate medical students were recruited with 85 and 75 responses collected, respectively. With one month of TBL experience prior to the study, the first-year students were now referred as the “novice” group. Second-year students had one year of TBL experience and were referred as the “experienced” group. Their age ranged from 18-23 (M=19.5, SD=1.06), with 103 (64%) males and 57 females.

### Assessing cognitive engagement

We used two validated self-reporting instruments to assess TBL engagement.

Firstly, the Situational Cognitive Engagement Measure (SCEM) was used to assess cognitive engagement at various TBL phases[15] (see Supplementary Digital Appendix 2 for complete survey). This 5-point Likert scale 4-question survey captures cognitive engagement of the ongoing activity at that instance. This tool developed items to assess the dynamic aspect of engagement, student’s will to persevere on the activity and how immersed they are which links back to the concepts of cognitive engagement. Previous studies[15] attained adequate reliability, including one in a similar local context[14].

Secondly, the Classroom Engagement Survey (CES) was used when each TBL session ended. This 5-point Likert scale 8-question survey retrospectively assesses overall engagement of the class (see Supplementary Digital Appendix 3 for complete survey). Its construct validity was established from previous TBL studies involving undergraduate nursing and medical students[30-32], obtaining high reliability with Cronbach Alpha of 0.80-0.89.

We decided to use both the SCEM and CES as measures of engagement as both instruments measure slightly different aspects of engagement. The SCEM measures how students perceive their immediate engagement while engaged with each TBL activity, while the CES asks students to retrospectively report how engaged they were throughout the TBL session. Cronbach’s alpha was calculated for internal reliability with values greater than 0.80 being desirable[37]. For the SCEM and CES instruments, our mean values were 0.72 and 0.83 respectively, indicating an acceptable to desirable degree of internal reliability.

Lastly, open-ended response questions were provided to further qualitatively explore students’ preferences with seating arrangement and engagement. Students were asked if they

(1) preferred sitting nearer to the tutor, (2) preferred sitting with their front-facing the tutor, (3) reasons for their seating preferences, and (4) if they preferred their original or new seating position.

To compare pre- and post-swap groups, independent-samples T-test was conducted to assess both the effects of seating distance and seating orientation on engagement. Significance levels were set at p<0.05. We used SPSS version 24 (IBM, Armonk, New York).

## Results

While the SCEM and CES tend to measure slightly different aspects of cognitive engagement, we found moderate correlation[38] between the two instruments in our study with Pearson’s Correlation r=0.57, p<.001.

### Seating distance and engagement

#### SCEM

In the novice cohort, Group A (students moving nearer to the tutor), there was no statistically significant change in SCEM scores. For Group B (those who moved further away), SCEM scores decreased significantly. Overall, between the pre-and post-session, SCEM scores decreased significantly for the entire cohort.

In the experienced cohort, there was no statistically significant change in SCEM scores for both Group A and Group B. Overall, between the pre-and post-sessions, there was no significant change in the SCEM scores. (Table 1)

**Table 1:**
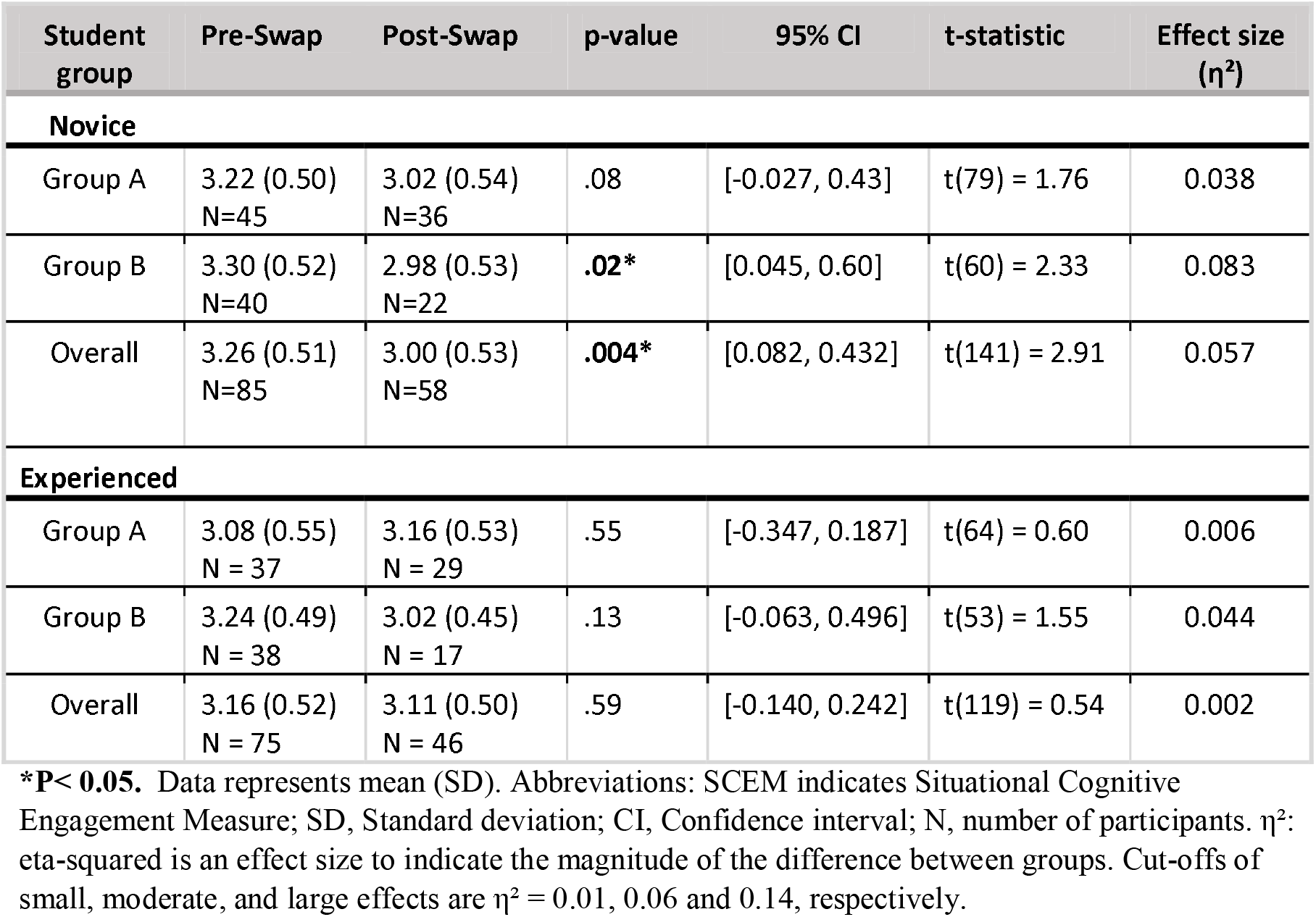
Mean SCEM Scores (SD) in Novice and Experienced Group Students Before and After Swapping Seating Arrangements at the Lee Kong Chian School of Medicine, Nanyang Technological University, Singapore.

#### CES

The results from the CES indicated a similar pattern to the SCEM results.

In the novice cohort, for Group A (students moving nearer to the tutor), CES scores did not significantly change while in Group B (those who moved further away), CES scores decreased significantly. Overall CES there was no statistically significant change in the pre- and post-session.

In the experienced cohort, for both Group A and B, CES scores were not statistically significant. Overall CES scores decreased slightly post-swap but were not statistically significant. (Table 2)

**Table 2:** Mean CES Scores in Novice and Experienced Group Students Before and After Swapping Seating Arrangements at the Lee Kong Chian School of Medicine, Nanyang Technological University, Singapore.

| Student group | Pre-Swap | Post-Swap | p-value | 95% CI | t-statistic | Effect size ($\eta^2$) |
| --- | --- | --- | --- | --- | --- | --- |
| **Novice** |  |  |  |  |  |  |
| Group A | 3.24 (0.66)<br>N = 45 | 3.23 (0.65)<br>N = 30 | .95 | [-0.298, 0.317] | t(73) = 0.06 | <0.001 |
| Group B | 3.38 (0.64)<br>N = 40 | 2.91 (0.63)<br>N = 20 | **.009\*** | [0.123, 0.820] | t(58) = 2.71 | 0.11 |
| Overall | 3.30 (0.65)<br>N = 85 | 3.10 (0.65)<br>N = 50 | .08 | [-0.025, 0.434] | t(133) = 1.76 | 0.023 |
| **Experienced** |  |  |  |  |  |  |
| Group A | 3.43 (0.53)<br>N = 37 | 3.41 (0.50)<br>N = 29 | .89 | [-0.240, 0.277] | t(64) = 0.14 | <0.001 |
| Group B | 3.62 (0.40)<br>N = 38 | 3.38 (0.60)<br>N = 17 | .08 | [-0.029, 0.522] | t(53) = 1.80 | 0.057 |
| Overall | 3.53 (0.47)<br>N = 75 | 3.40 (0.53)<br>N = 46 | .17 | [-0.057, 0.315] | t(119) = 1.38 | 0.016 |

**\*P < 0.05.** Abbreviations: CES indicates Classroom Engagement Survey; SD, Standard deviation; CI, Confidence interval; N, number of participants. $\eta^2$ : eta-squared is an effect size to indicate the magnitude of the difference between groups. Cut-offs of small, moderate, and large effects are $\eta^2 = 0.01, 0.06$ and $0.14$ , respectively.

### Seating orientation and engagement

In both novice and experienced students, SCEM scores for those with their fronts facing the tutor were lower than those with their backs facing the tutor but were not statistically significant.

In novice students, CES scores for those with their fronts facing the tutor was higher than those with their backs facing the tutor but was not statistically significant. In experienced students, CES scores for those with their fronts facing the tutor was lower than those with their backs facing the tutor but was not statistically significant. (Table 3)

**Table 3:**
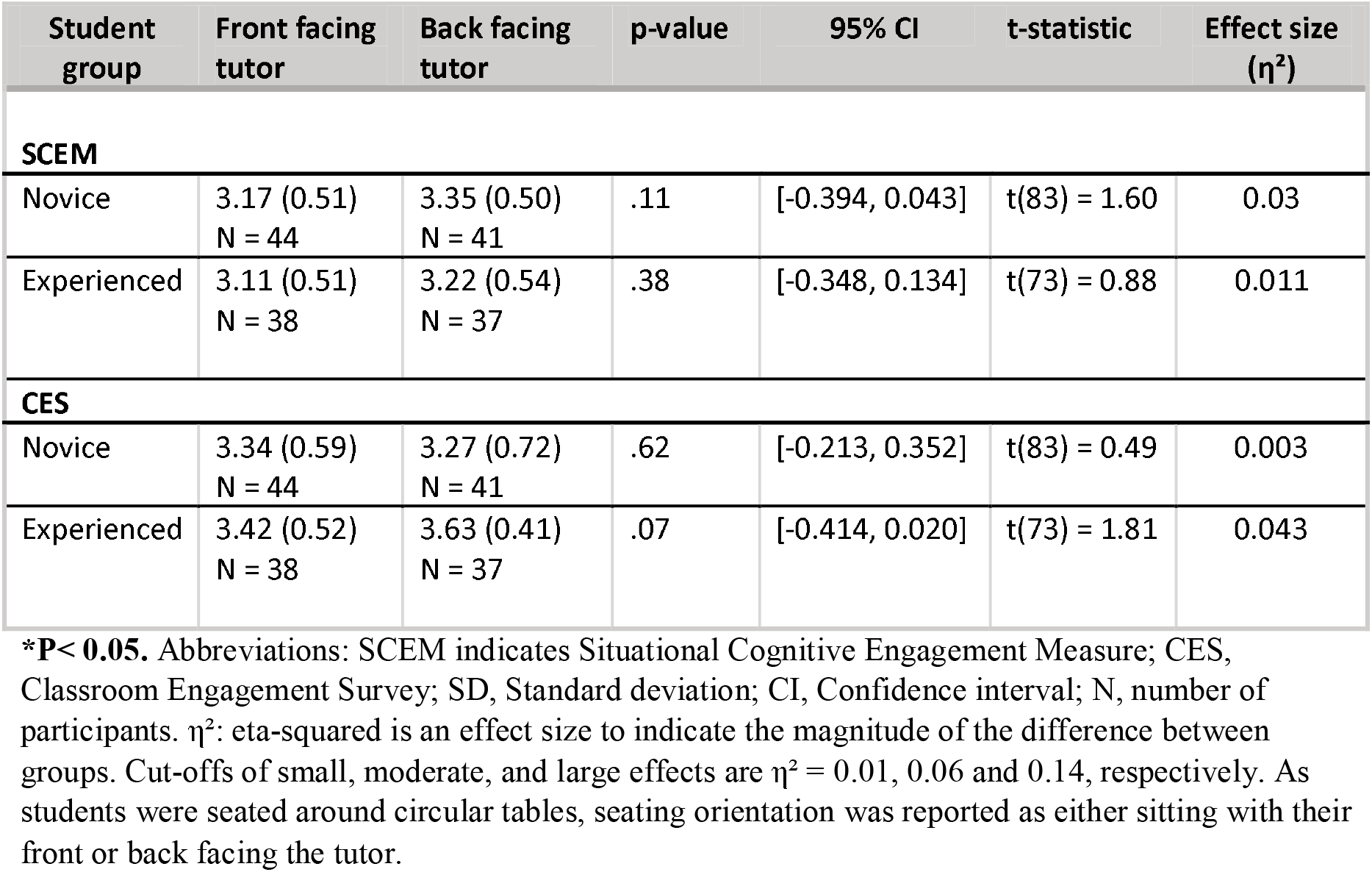
Comparing Seating Orientation and Mean SCEM and CES Scores in Novice and Experienced Students at the Lee Kong Chian School of Medicine, Nanyang Technological University, Singapore.

### Interaction between seating distance and orientation

A two-way analysis of variance (ANOVA) was performed to assess the interaction between seating distance and orientation which was not statistically significant. Novice group: F(1,81)=0.228, p=0.63. Experienced group: F(1,71)=0.937, p=0.34.

### Student preferences

Participants were asked to rate, on a scale of 1 (strongly disagree) to 5 (strongly agree), how much they preferred sitting *nearer* to the tutor and sitting with their front-*facing* the tutor.

- Novice students preferred sitting nearer to the tutor compared to sitting further away. (M=3.45 vs M=2.53) (p<0.001) while experienced students had no statistically significant difference in such preferences (M=2.62 vs M=2.61) (p=0.95).
- Both novice and experienced students had a statistically significant preference to sit with their front-facing the tutor compared to facing the back. Novice: M=4.20 vs M=3.56 (p=0.001). Experienced: M=4.21 vs M=3.62 (p=0.003)

Further analysis revealed that 86 (51%) students felt that seating distance did not affect their learning, with common reasons such as screens being all around the room and audio from the speaker being well heard. 104 (61%) preferred sitting with their front-facing the tutor, with common reasons being a preference to look at the tutor while they are speaking and feeling more focused. Some quotes from students include: “I don’t think it (seating position) affects my learning as there are **screens around the studio**”, “The **classroom** is **360**° so it doesn’t matter”, “**audio** from the front of the class can be **easily heard** everywhere” and “**acoustics** are good enough such that the lesson can be heard clearly”.

## Discussion

We investigated the effects of seating distance and orientation to the tutor on TBL engagement in a large classroom specifically designed for TBL. Engagement levels were assessed before and after seating positions were swapped, using two separate instruments.

Novice students showed a significant drop in engagement in those that moved further with a moderate effect size which supports the original hypothesis. However, when analysing them together, overall engagement dropped post-swap regardless of direction moved. Experienced students showed no significant change in engagement based on seating distance.

Additionally, novice students displayed a stronger preference to sit nearer to the tutor compared to experienced students.

From these, it is highly likely that the very act of changing seating layouts influenced the novice groups’ engagement. On closer analysis of students’ preferences, we asked if they preferred their original or new seats and found that 40 (80%) novice students preferred their original seats compared to only 21 (48%) experienced students. A possible explanation is that the novice group, being only one month into medical school, were inexperienced with TBL pedagogy and still familiarising with learning in a new university. After adjusting into this new learning style for one month, any change could influence how they pay attention in class. Experienced students had at least one-year of TBL experience and were well acclimatised to learning in such settings. A simple change in seating distance hardly affected their engagement. This may illustrate that as students become more familiar with the TBL process, they acquire skills which compensate for any minor distractions caused by the physical environment. Novice students were more susceptible to changes in the physical environment and may have a higher need for psychological safety.

For both groups, engagement levels were not significantly impacted by seating orientation. But they strongly preferred to sit with their front-facing the tutor. From general classroom observations, some students compensated by turning their whole body to face the tutor. With circular tables and a decentralised classroom, it is impossible for every student to sit with their front-facing the tutor. While the audio-visual system allows students to follow discussions even when seated far back, it cannot compensate for students’ preference to look at the tutor.

Overall, our results refute the initial hypotheses that sitting nearer or facing the tutor results in higher engagement because students were not significantly affected by their seating arrangement.

### Practical implications

The findings of this paper provide important and practical implications on seating arrangement in the TBL classroom. Juxtaposing the strong preference to sit nearer or face the tutor and the insignificant effect of seating arrangement on engagement, there appear to be deeper factors at play. While students displayed a preference to sit nearer or face the tutor, they can overcome such physical inconveniences and feel similarly engaged at their less ideal spot. We postulate that medical students generally have higher motivation levels given the work needed to get into medical school and the commitment to a specific career[39]. With this motivation to learn, they adapt to wherever they seat and pay attention regardless. Also, tutors are relatively less involved compared to team interactions for the overall TBL experience (assessed with CES tool), possibly explaining why students were not affected by seating arrangement and reinforce the rationale of TBL as a learning modality.

Quotes from students showed that classroom design was influential for their learning. The implication is that learning space design does have an impact on learning and engagement. While other institutions are compelled to use traditional learning spaces for TBL, our institution specially designed the classroom, according to well-documented architectural and human factors, to facilitate TBL[26]. The circular layout with dual tiered seating, six screens and microphones at every table allows students to follow and contribute to discussions wherever they are. Learning spaces are only as successful as the students’ motivation and willingness to best make use of it[40]. This motivation allows students to fully utilise the technological space and adapt to less preferred seating positions.

Other studies comparing traditional and active learning classrooms substantiates this, showing improved ease of working in teams[33], higher student engagement[41, 42] and grades[43, 44] in specialised classrooms for interactive pedagogies. Previous reports highlighted that aligning physical learning spaces with the curriculum is valuable to the holistic health professions education[3]. Classroom layouts that allowed students to see everyone contributed to the perception of a safe learning space[45]. Therefore, such team- centric learning spaces provide an engaging setting, favourable for active, collaborative learning[26].

## Limitations

This study has some limitations. Firstly, as all TBL sessions differed, results may be confounded by factors such as TBL topics, TBL length, student fatigue and tutor variation. This was especially true in novice students where post-swap engagement dropped significantly as highlighted above. Secondly, only medical students were recruited and are not representative of the general TBL population, which includes other health professions[22], science[24, 46] and business[47] courses. Thirdly, due to sample size constraints, this study had no control group that did not change seats. Changing seats itself may be disruptive and having a group which swapped seats but not move nearer or further would make comparisons more robust. Lastly, survey responses fell significantly after the swap, with 64 (40%) fewer responses from 160 to 96, which may affect the accuracy and reliability of the results. Possible reasons include the interruptive nature of the survey as lesson flow was disrupted three times. But this was unavoidable to capture ongoing engagement. Given these limitations, this study contributes meaningfully to the literature by being the first quasi-experimental study that we know of to explore the relationship between seating arrangement and engagement in TBL.

## Conclusion

There were no significant effects of seating distance or orientation on TBL engagement. There was, however, a preference for novice students to sit nearer to the tutor. Also, both experienced and novice students had a strong preference to sit with their fronts facing the tutor. Compared to experienced students, novice students were affected more by changes in seating arrangement who may showcase their need for early psychological safety.

The specially designed classroom were important reasons students felt unaffected by seating arrangements. The findings reassure us that, with proper attention to the physical space, students are not at a disadvantage from their seating arrangement, which supports utilising specialised active learning classrooms for TBL.

## Data Availability

All relevant data referred to is available in the tables of the manuscript. A full data set is in the possession of the authors.

## Acknowledgements

The authors wish to thank the undergraduate students from LKCMedicine Class of 2023 and 2024 for their kind participation in the surveys.

## Ethical Approval

The study was approved by the Institutional Review Board of Nanyang Technological University [2019-06-036].

## Supplementary Appendices

### Supplementary Appendix 1

Topics and questions for the 4 TBL sessions

First-year students’ session 1 pre-swap

|  |  |
| --- | --- |
| Topic | Foundations of Medicine (Cell division, Cell signalling)<br>The whole TBL session from readiness assurance to AE lasted from 9am to 6pm. BQ and AE phase lasted from 1300 to 1730. |
| BQ | Survey was filled in 15-30mins into BQ phase. BQ phase lasted 2hour.<br>BQ Qn: DNA replication involves a proofreading function to maintain a very low rate of error frequency. What prevents the old DNA strand from being "edited"? |
|  | 1. **Methyl groups**<br>2. RNA primers<br>3. Okazaki fragments<br>4. Nucleoside analogues |
| AE | AE was filled in 45mins into AE phase. Total AE phase lasted 2 ½hour.<br>• AE1: In the following genomic DNA seq, the start codon is underlined: 5'-TTAGCTGTGTAG<u>ATG</u>CGGCATGCAT. After exposing cells to irradiation, an insertion of adenosine was identified 4 nucleotides before the start codon. The new sequence is: 5'-TTAGCTGTGATAG<u>ATG</u>CGGCATGCAT. What are the potential consequences of this event? One or more answer may be correct (2marks)<br>1. The insertion may induce a shift in the reading frame<br>2. **The insertion may affect transcription of this gene**<br>3. The insertion may affect posttranslational modifications<br>4. The insertion may lead to a truncated protein<br>5. **The insertion may not have any impact**<br>• AE2: List some advantages and disadvantage of molecular techniques (qpCR, Northern blot, microarray, RNA seq) |

First-year students’ session 2 post-swap

First-year students' session 2 post-swap
|  |  |
| --- | --- |
| Topic: | Foundations of Medicine (Anatomy, Histology, Embryology).<br>BQ phase lasted from 1030 to 12nn. AE phase lasted from 1400 to 1545 – 3hr 15mins |
| BQ | Survey was filled in 25mins into BQ<br>• BQ Qn: A 5-day-old baby was presented to the clinic by his parents with a 10cm mass the lower back (sacroccygeal region) since birth. He was delivered at home |
|  | traditional birth attendants after a term, unsupervised pregnancy to a 32-year old woman. Clinical and radiological examination confirms the diagnosis of sacrococcygeal teratoma. Which of the following gives rise to this type of tumour?<br>1. Neural crest<br>2. Neural tube<br>3. Remnants of notochord<br>4. Remnants of primitive streak<br>5. Splanchnic mesoderm |
| AE | Survey was filled in 30mins into AE phase<br>• AE1: Endocytosis is a process which brings extracellular material into the cell's cytoplasm. During endocytosis of LDL, which of the following does not occur?<br>1. Formation of clathrin-coated pits<br>2. Incorporation of membrane receptors into clathrin-coated pits<br>3. Fusion of endosome and formation of lysosome<br>4. Release of cholesterol into cytoplasm from lysosome<br>5. Excretion of protons from endosome<br>Rationale: Protons enter endosome (they are not excreted) which becomes the lysosome where protons (low pH) activate pH-sensitive hydrolytic enzymes which degrade contents. |

Second-year students’ session 1 pre-swap

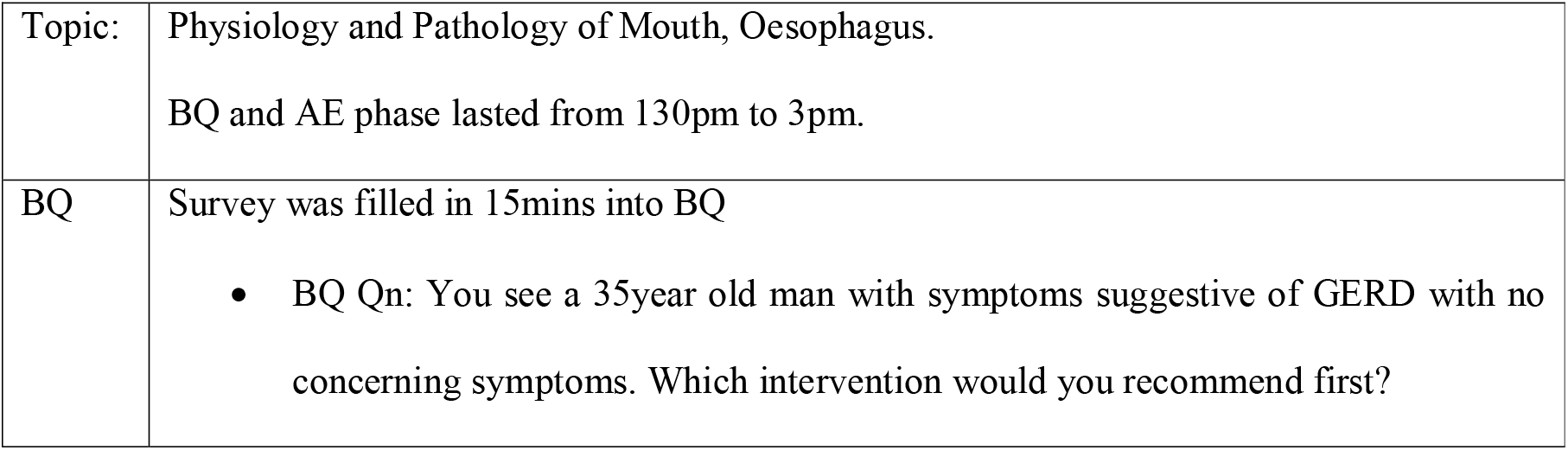
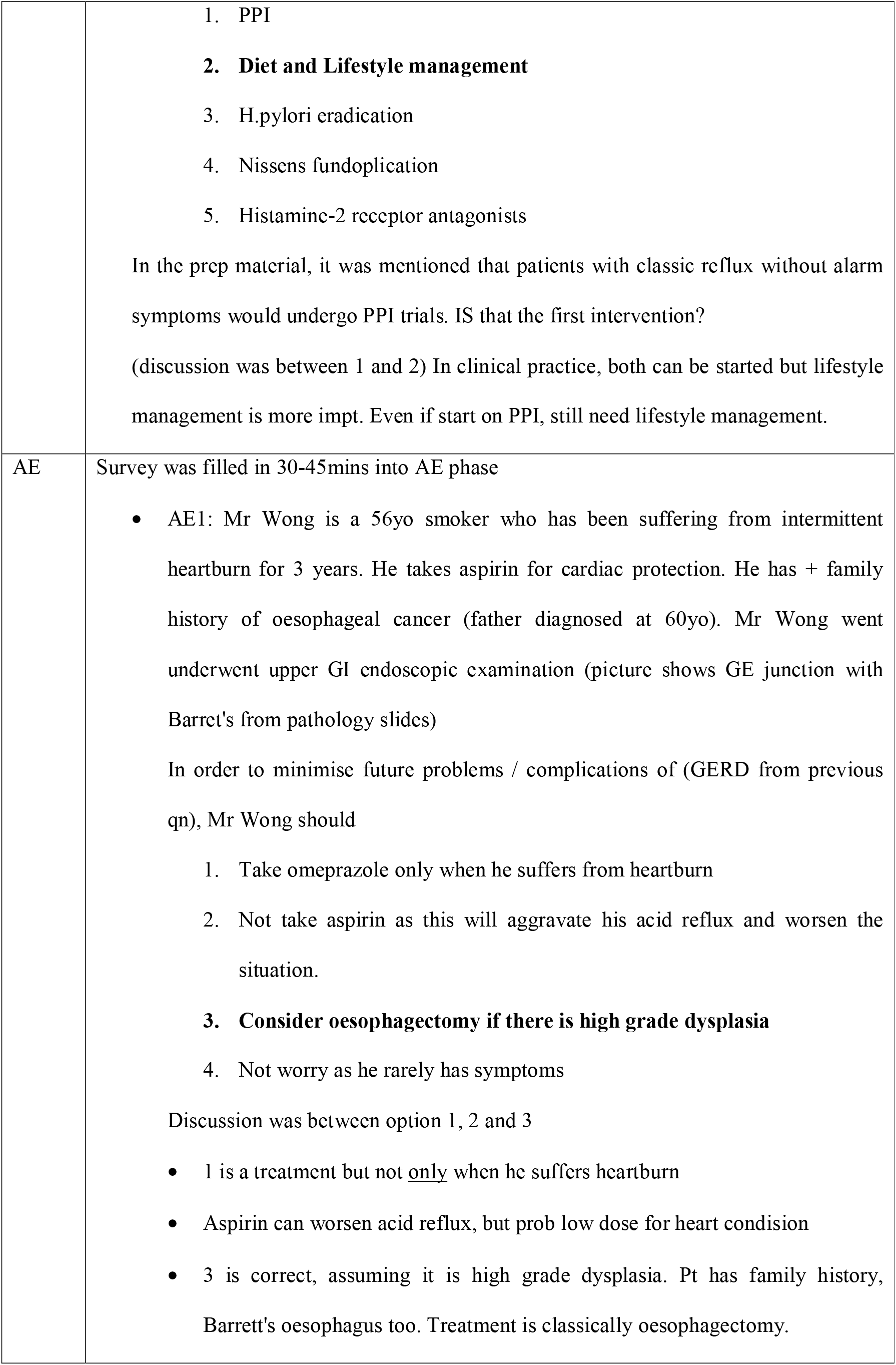
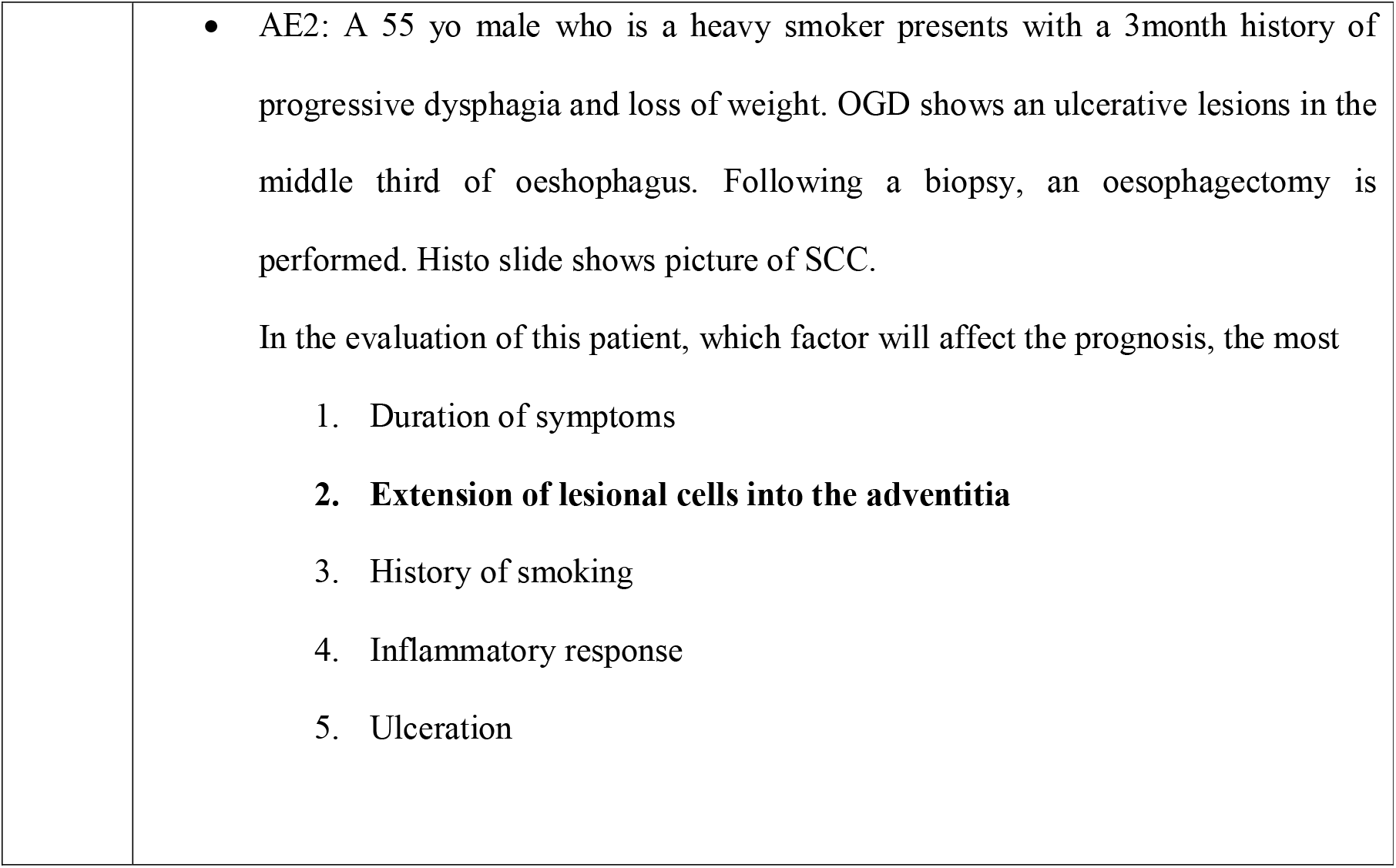

Second-year students' session 1 pre-swap
|  |  |
| --- | --- |
| Topic: | Physiology and Pathology of Mouth, Oesophagus.<br>BQ and AE phase lasted from 130pm to 3pm. |
| BQ | Survey was filled in 15mins into BQ<br>• BQ Qn: You see a 35-year old man with symptoms suggestive of GERD with no concerning symptoms. Which intervention would you recommend first? |
|  | 1. PPI<br>**2. Diet and Lifestyle management**<br>3. H.pylori eradication<br>4. Nissens fundoplication<br>5. Histamine-2 receptor antagonists<br>In the prep material, it was mentioned that patients with classic reflux without alarm symptoms would undergo PPI trials. IS that the first intervention?<br>(discussion was between 1 and 2) In clinical practice, both can be started but lifestyle management is more imp. Even if start on PPI, still need lifestyle management. |
| AE | Survey was filled in 30-45mins into AE phase<br>• AE1: Mr Wong is a 56yo smoker who has been suffering from intermittent heartburn for 3 years. He takes aspirin for cardiac protection. He has + family history of oesophageal cancer (father diagnosed at 60yo). Mr Wong went underwent upper GI endoscopic examination (picture shows GE junction with Barret's from pathology slides)<br>In order to minimise future problems / complications of (GERD from previous qn), Mr Wong should<br>1. Take omeprazole only when he suffers from heartburn<br>2. Not take aspirin as this will aggravate his acid reflux and worsen the situation.<br>**3. Consider oesophagectomy if there is high grade dysplasia**<br>4. Not worry as he rarely has symptoms<br>Discussion was between option 1, 2 and 3<br>• 1 is a treatment but not <u>only</u> when he suffers heartburn<br>• Aspirin can worsen acid reflux, but prob low dose for heart condition<br>• 3 is correct, assuming it is high grade dysplasia. Pt has family history, Barrett's oesophagus too. Treatment is classically oesophagectomy. |
|  | • AE2: A 55 yo male who is a heavy smoker presents with a 3month history of progressive dysphagia and loss of weight. OGD shows an ulcerative lesions in the middle third of oesophagus. Following a biopsy, an oesophagectomy is performed. Histo slide shows picture of SCC.<br>In the evaluation of this patient, which factor will affect the prognosis, the most<br>1. Duration of symptoms<br>2. **Extension of lesional cells into the adventitia**<br>3. History of smoking<br>4. Inflammatory response<br>5. Ulceration |

Second-year students’ session 2 post-swap

Second-year students' session 2 post-swap
|  |  |
| --- | --- |
| Topic | Physiology and Pathology of Stomach<br>BQ and AE phase lasted from 130pm to 250pm |
| BQ | **BQ** (lasted from 135pm to 2pm). Survey was given 25mins into BQ<br>• BQ Qn: Risk factors for gastric cancer?<br>1. Zollinger Ellison syndrome<br>2. **Pernicious anaemia**<br>3. Patients >75<br>4. Patients with Barrett's oesophagus<br>Discussion was on option 1: Why is Zollinger Ellison syndrome wrong |
| AE | **AE** (Lasted from 2pm to 250pm). Survey was given 15 mins into AE.<br>• AE1: Which of the following investigations will contribute significantly towards treatment of MR Tan's problems?<br>1. Serum gastrin levels |
|  | 2. Serum H. pylori antibody<br>**3. Urea breathe test**<br>4. Stool H pylori antigen test<br>A staging CT scan suggests a very large gastric cancer in the remnant stomach with suggestion of liver metastasis. Palliative chemotherapy was recommended<br>• AE2: In what circumstances would surgery be indicated in this patient? (eg advanced gastric cancer)<br>1. Persistent epigastric pain<br>2. Unable to tolerate full meal, and intermittent vomiting<br>3. Intermittent melaena and blood in vomitus<br>**4. Fever, abdominal pain and guarding**<br>Option 4 implies perforation and peritonitis. |

### Supplementary Appendix 2

Example of the Situational Cognitive Engagement Measure.

It consists of 4 questions on a 5-point Likert scale. During the Burning Questions / Application Exercise, on a scale from 1 to 5, with 1 (not true at all) and 5 (very true for me), please indicate how true the statements are for you:

1. I am engaged with the topic at hand
2. I put in a lot of effort understanding the topic
3. I wish I could still continue for a while
4. I am so involved that I forget everything around me

### Supplementary Appendix 3

Example of the Classroom Engagement Survey (CES).

It consists of 8 questions on a 5-point Likert scale with 1 (Strongly disagree) and 5 (Strongly agree). From the following statements, please choose the option that best described the extent to which you **agree** with about **today’s** class.

1. Most students were actively involved
2. I had fun in class today
3. I contributed meaningfully to class discussions
4. Most students were not paying attention
5. I paid attention most of the time
6. I did not enjoy class today
7. I participated in the class most of the time
8. I would like more class sessions to be like this one

## References

1. Beichner RJ. History and Evolution of Active Learning Spaces. New Directions for Teaching and Learning. 2014;2014(137):9–16.

2. Kolb AY, Kolb DA. Learning Styles and Learning Spaces: Enhancing Experiential Learning in Higher Education. Academy of Management Learning & Education. 2005;4(2):193–212.

3. Nordquist J, Sundberg K, Laing A. Aligning physical learning spaces with the curriculum: AMEE Guide No. 107. Medical Teacher. 2016;38(8):755–68.

4. Oblinger DG. Learning Spaces: Educase; 2006. Available from: https://www.educause.edu/ir/library/pdf/PUB7102.pdf.

5. O’Hare M. Classroom Design for Discussion-Based Teaching. Journal of Policy Analysis and Management. 1998;17(4):706–20.

6. Shernoff DJ, Sannella AJ, Schorr RY, et al. Separate worlds: The influence of seating location on student engagement, classroom experience, and performance in the large university lecture hall. Journal of Environmental Psychology. 2017;49:55–64.

7. Becker FD, Sommer R, Bee J, Oxley B. College Classroom Ecology. Sociometry. 1973;36(4):514–25.

8. Ridling Z. The Effects of Three Seating Arrangements on Teachers’ Use of Selective Interactive Verbal Behaviors. Paper presented at the Annual Meeting of the American Educational Research Association New Orleans, LA. 1994.

9. Marx A, Fuhrer U, Hartig T. Effects of Classroom Seating Arrangements on Children’s question-asking. Learning Environments Research. 1999;2(3):249–63.

10. Appleton JJ, Christenson SL, Kim D, Reschly AL. Measuring cognitive and psychological engagement: Validation of the Student Engagement Instrument. Journal of School Psychology. 2006;44(5):427–45.

11. Fredricks JA, Blumenfeld PC, Paris AH. School Engagement: Potential of the Concept, State of the Evidence. Review of Educational Research. 2004;74(1):59–109.

12. Richardson JC, Newby T. The Role of Students’ Cognitive Engagement in Online Learning. American Journal of Distance Education. 2006;20(1):23–37.

13. Walker CO, Greene BA, Mansell RA. Identification with academics, intrinsic/extrinsic motivation, and self-efficacy as predictors of cognitive engagement. Learning and Individual Differences. 2006;16(1):1–12.

14. Rotgans JI, Schmidt HG, Rajalingam P, et al. How cognitive engagement fluctuates during a team-based learning session and how it predicts academic achievement. Advances in Health Sciences Education. 2018;23(2):339–51.

15. Rotgans JI, Schmidt HG. Cognitive engagement in the problem-based learning classroom. Advances in Health Sciences Education. 2011;16(4):465–79.

16. Freeman S, Eddy SL, McDonough M, et al. Active learning increases student performance in science, engineering, and mathematics. Proceedings of the National Academy of Sciences. 2014;111(23):8410–5.

17. Sinatra GM, Heddy BC, Lombardi D. The Challenges of Defining and Measuring Student Engagement in Science. Educational Psychologist. 2015;50(1):1–13.

18. Springer L, Stanne ME, Donovan SS. Effects of Small-Group Learning on Undergraduates in Science, Mathematics, Engineering, and Technology: A Meta-Analysis. Review of Educational Research. 1999;69(1):21–51.

19. Michaelsen LK, Knight AB, Fink LD. Team-based learning : a transformative use of small groups in college teaching. Sterling, VA: Stylus Pub.; 2004.

20. Koles PG, Stolfi A, Borges NJ, Nelson S, Parmelee DX. The Impact of Team-Based Learning on Medical Students’ Academic Performance. Academic Medicine. 2010;85(11):1739–45.

21. Parmelee DX, Hudes P. Team-based learning: A relevant strategy in health professionals’ education. Medical Teacher. 2012;34(5):411–3.

22. Reimschisel T, Herring AL, Huang J, Minor TJ. A systematic review of the published literature on team-based learning in health professions education. Medical Teacher. 2017;39(12):1227–37.

23. Currey J, Eustace P, Oldland E, Glanville D, Story I. Developing professional attributes in critical care nurses using Team-Based Learning. Nurse Education in Practice. 2015;15(3):232–8.

24. Zgheib NK, Dimassi Z, Bou Akl I, Badr KF, Sabra R. The long-term impact of team-based learning on medical students’ team performance scores and on their peer evaluation scores. Medical Teacher. 2016;38(10):1017–24.

25. Parmelee DX, Michaelsen LK. Twelve tips for doing effective Team-Based Learning (TBL). Medical Teacher. 2010;32(2):118–22.

26. Rajalingam P, Rotgans JI, Zary N, Ferenczi MA, Gagnon P, Low-Beer N. Implementation of team-based learning on a large scale: Three factors to keep in mind*. Medical Teacher. 2018;40(6):582–8.

27. Hrynchak P, Batty H. The educational theory basis of team-based learning. Medical Teacher. 2012;34(10):796–801.

28. Schmidt HG, Rotgans JI, Rajalingam P, Low-Beer N. Knowledge ReconsolidPublish Ahead of Print.

29. Kelly PA, Haidet P, Schneider V, Searle N, Seidel CL, Richards BF. A Comparison of In-Class Learner Engagement Across Lecture, Problem-Based Learning, and Team Learning Using the STROBE Classroom Observation Tool. Teaching and Learning in Medicine. 2005;17(2):112–8.

30. Clark MC, Nguyen HT, Bray C, Levine RE. Team-based learning in an undergraduate nursing course. The Journal of nursing education. 2008;47(3):111–7.

31. Levine RE, O’Boyle M, Haidet P, et al. Transforming a Clinical Clerkship with Team Learning. Teaching and Learning in Medicine. 2004;16(3):270–5.

32. Mennenga HA. Team-based learning: Engagement and accountability with psychometric analysis of a new instrument. UNLV Theses, Dissertations, Professional Papers, and Capstones. 2010.

33. Espey M. Does Space Matter? Classroom Design and Team-Based Learning. Applied Economic Perspectives and Policy. 2008;30(4):764–75.

34. Hong JM, Rajalingam P. Geographic trends in Team-Based Learning (TBL) research and implementation in medical schools. Health Professions Education. 2019.

35. Zomorodian K, Parva M, Ahrari I, et al. The effect of seating preferences of the medical students on educational achievement. Medical Education Online. 2012;17(1):10448.

36. Parmelee DX, Michaelsen LK, Cook S, Hudes PD. Team-based learning: A practical guide: AMEE Guide No. 65. Medical Teacher. 2012;34(5):e275–e87.

37. Polit DF, Beck CT. Nursing Research: Generating and Assessing Evidence for Nursing Practice. 8th Edition. New York: Lippincott, Williams, & Wilkins.; 2008.

38. Akoglu H. User’s guide to correlation coefficients. Turk J Emerg Med. 2018;18(3):91–3.

39. Kusurkar RA, Ten Cate TJ, van Asperen M, Croiset G. Motivation as an independent and a dependent variable in medical education: a review of the literature. Med Teach. 2011;33(5):e242–62.

40. Nordquist J, Laing A. Designing spaces for the networked learning landscape. Medical Teacher. 2015;37(4):337–43.

41. Whiteside A, Brooks D, Walker J. Making the Case for Space: Three Years of Empirical Research on Learning Environments. Educase Quarterly. 2010;33.

42. Sanders MJ. Classroom Design and Student Engagement. Proceedings of the Human Factors and Ergonomics Society Annual Meeting. 2013;57(1):496–500.

43. Brooks DC. Space matters: The impact of formal learning environments on student learning. British Journal of Educational Technology. 2011;42(5):719–26.

44. Cotner S, Loper J, Walker J, Brooks D. Research and Teaching: “It’s Not You, It’s the Room”--Are the High-Tech, Active Learning Classrooms Worth It? Journal of College Science Teaching. 2013;042.

45. Holley LC, Steiner S. SAFE SPACE: STUDENT PERSPECTIVES ON CLASSROOM ENVIRONMENT. Journal of Social Work Education. 2005;41(1):49–64.

46. Yuretich RF, Kanner LC. Examining the Effectiveness of Team-Based Learning (TBL) in Different Classroom Settings. Journal of Geoscience Education. 2015;63(2):147–56.

47. Sisk RJ. Team-based learning: systematic research review. The Journal of nursing education. 2011;50(12):665–9.

